# Analysis of the specificity of the SD Biosensor Standard Q Ag-Test based on Slovak mass testing data

**DOI:** 10.1101/2020.12.08.20246090

**Authors:** Michal Hledík, Jitka Polechová, Mathias Beiglböck, Anna Nele Herdina, Robert Strassl, Martin Posch

## Abstract

From 31.10. - 1.11.2020 Slovakia has used the SD Biosensor Standard Q Ag-Test for nationwide tests for SARS-CoV-2, in which 3,625,332 persons from 79 counties were tested. Based on this data, we calculate that the specificity of the test is at least 99.6% (with a 97.5% one-sided lower confidence bound). Our analysis is based on a worst case approach in which all positives are assumed to be false positives. Therefore, the actual specificity is expected to exceed 99.6%.

## Aim

While PCR-tests are usually considered as the gold standard to detect infection with the SARS-CoV-2 coronavirus in terms of sensitivity as well as specificity, Antigen-Tests (Ag-Tests) offer practical advantages in terms of costs, logistics and speed [1]. Because Ag-Tests may play a major role in large scale testing strategies [2,3], their specificity is of significant interest.

In this note we consider the specificity of the SD Biosensor Standard Q Ag-Test based on data of mass tests in Slovakia and infer Ag-Test specificity from a large sample of the general population. This is in contrast to other studies which use a PCR-test as a reference to estimate sensitivity and specificity of an Ag-test [4-7].

## Data structure and collection

From late October to early November 2020, Slovakia undertook large scale testing of its population [8]. To the best of our knowledge exclusively the SD Biosensor Standard Q Ag-Test was used [9].

The tests in Slovakia were divided into three phases. In the first pilot phase, testing was only conducted in certain particularly affected counties. In phase 2 (31.10. - 1.11.) all Slovak counties were tested. In phase 3 (6.11. - 8.11.) all heavily affected counties (those with > 0.7% prevalence during phase 2) were tested again. In this retrospective study, we use publicly available data on the outcome of tests in phase 2 [10].

In phase 2, all residents aged 10 to 65 throughout Slovakia were invited to get tested and 3 625 332 participated (about 66% of the population of Slovakia). Among 79 administrative counties of Slovakia, participation rate varied from 39% (Košice III county) to 78% (Senec county) of inhabitants. Participation was voluntary, but it was a condition to avoid quarantine. Persons that were quarantined due to a previous positive PCR-test for COVID-19 or due to being a close contact of such a person were excluded from the test. Data from phase 2 was used in this study, because the general countrywide testing, performed irrespective of regional incidence rates, potentially covers also regions with very low incidence. Results from the latter are most suitable to derive bounds of the specificity.

Participants were tested with antigen tests at specially set up locations by medical personnel via nasopharyngeal swab sampling and received their results after a short waiting period.

## Statistical analysis

To derive a lower bound for the specificity we made the conservative (“worst case”) assumption that potentially all positive results constitute false positive results. For each of the 79 counties, we compute the rate of positive tests together with two-sided binomial Clopper-Pearson confidence intervals at the Bonferroni-adjusted significance level of alpha = 0.05/79 (adjusted for the number of counties; see e.g. [11]). The minimum upper bound of these adjusted confidence intervals is an upper 97.5% confidence bound for the test positivity rate. This will be in a county with low disease activity and a large sampled population. Under the conservative assumption that all positive results constitute false positives, it is also an upper bound on the false positive rate of the test and defines a lower bound for the true specificity.

Figure 1 depicts the test positivity rate of the individual counties ordered according to the upper bound of the simultaneous Bonferroni adjusted 95% confidence interval from low to high incidence. The lowest upper bound (obtained for Rožnava) is 0.40%. In terms of specificity (instead of false positives) this implies that with (97.5% confidence) the specificity of Standard Q is higher than 99.6%.

**Figure 1:**
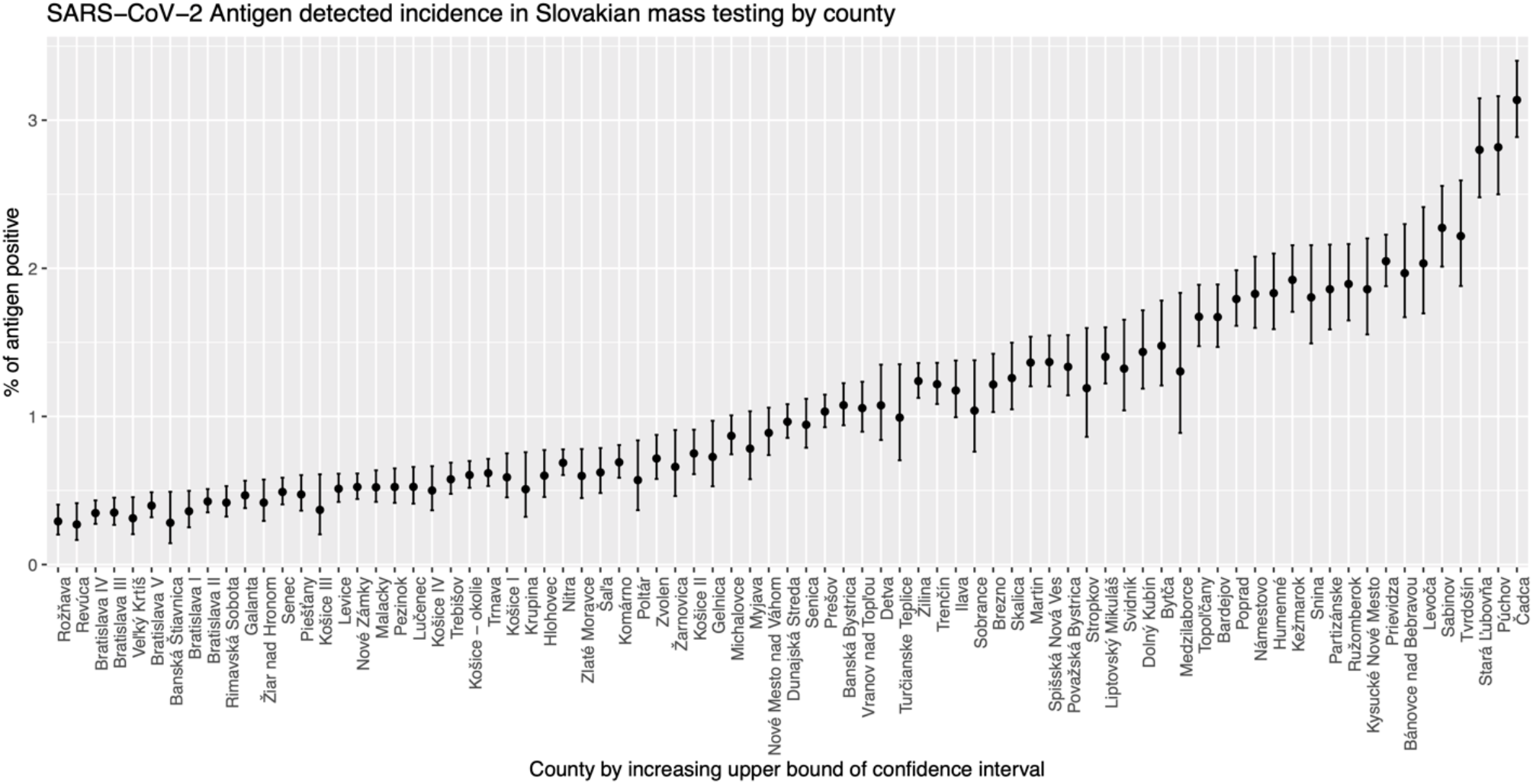
SARS-CoV-2 antigen detected incidence in Slovakian mass testing by county. Test positivity rates among the 3,625,332 tested persons in the 79 counties with simultaneous Bonferroni adjusted 95% confidence intervals. Counties are ordered from low to high incidence by the upper confidence bound. Data source [10].

Table 1 shows the five counties with the lowest upper bound on the positivity rate of antigen tests. These include counties with both relatively low participation rate (Rožňava, Revúca and Veľký Krtíš rank 65., 67. and 57. in participation among the 79 counties) and relatively high participation rate (Bratislava IV and Bratislava III rank 13. and 4.).

| County | Positivity rate upper bound simultaneous 95% CI | Number of tests | Number of positive tests | Participation, % tested of inhabitants | Number of inhabitants 2019 [12] |
| --- | --- | --- | --- | --- | --- |
| Rožňava | 0.40% | 34307 | 100 | 55% | 62131 |
| Revúca | 0.41% | 21419 | 58 | 54% | 39537 |
| Bratislava IV | 0.43% | 65861 | 229 | 67% | 97792 |
| Bratislava III | 0.45% | 49788 | 175 | 72% | 69479 |
| Veľký Krtíš | 0.46% | 24282 | 76 | 56% | 43263 |

## Discussion and conclusions

This worst case analysis provides only a lower bound of the true specificity since it neglects entirely the true incidence of COVID-19. As the antigen tests in Slovakia were not controlled directly using accompanying PCR-tests, we refrain from an attempt to subdivide the observed positives into true and false positives.

The binomial distribution assumption underlying the computation of the Clopper-Pearson confidence intervals, is based on the assumption of independent events that could be violated if, e.g. testing stations were operating with different quality. However, the Bonferroni-correction is a strictly conservative approach to derive simultaneous confidence bounds that account for the selection of the county with the lowest upper bound to derive the estimate.

The above estimate is consistent with the information provided by the manufacturer [13], stating 0.32% (0.01, 1.78) as a false positive rate - a very broad CI. The derived upper bound from the mass testing is more informative and appears relevant in that it constitutes the first (to the best of our knowledge) bound obtained from large scale practical use of Standard Q and also suggests (slightly) better performance of Standard Q compared to previous studies: [4], a large study (with 2347 SARS-CoV-2-free samples based on a PCR-test), suggests a specificity of 99.3% (CI 98.6-99.6). Other available specificity estimates are based on an order of magnitude smaller samples (99.2% (CI 97.1-99.8) [7] and 100% for [5]) and, in addition, are from study populations with high incidence rates (according to PCR-testing). [6] gives a point estimate of 98.53% based on 100 SARS-Cov2-free samples with other respiratory viruses present and 35 healthy volunteers.

It is well understood in the epidemiology literature that imperfect reference criteria lead to a systematic bias of the true incidence [14]. Specifically, the sensitivity and specificity of tests are systematically underestimated if the criterium (SARS-CoV-2 infection) cannot be assessed directly and tests are compared to a surrogate criterion as gold standard which is subject to errors. This could be a contributing factor for the relatively higher specificity found in Slovak data compared to other studies. Other conceivable factors that could bias the results include the different quality of swab sampling and handling, low temperature in outdoor testing stations and deviations in the production over time and data quality.

## Data Availability

All data are publicly available and referenced in the MS.

https://github.com/Institut-Zdravotnych-Analyz/covid19-data/blob/main/Slovakia_National_Testing_Municipality_Data.csv

## Acknowledgements

We would like to thank Alfred Uhl, Richard Kollár and Katarína Bod’ová for very helpful comments. We also thank Matej Mišík for discussion and information regarding the Slovak testing data and Ag-test used.

